# Predicting Relapse and Psychosocial Functioning in Major Depressive Disorder: A Machine Learning Approach Using Clinical and Resting-State fMRI Data

**DOI:** 10.1101/2025.10.22.25334871

**Authors:** Lena Marie Puder, Nils Ralf Winter, Janik Goltermann, Susanne Meinert, Kira Flinkenflügel, Judith Krieger, Julia Hubbert, Ramona Leenings, Lukas Fisch, Jan Ernsting, Carlotta Barkhau, Max Konowski, Dominik Grotegerd, Theresa Slump, Hannah Meinert, Elisabeth Schrammen, Luisa Altegoer, Elisabeth J. Leehr, Paula Usemann, Lea Teutenberg, Frederike Stein, Florian Thomas-Odenthal, Benjamin Straube, Nina Alexander, Hamidreza Jamalabadi, Andreas Jansen, Igor Nenadic, Tilo Kircher, Joachim Gross, Tim Hahn, Udo Dannlowski

## Abstract

**Background:** Machine learning approaches pave a promising avenue to advance individual predictions about psychiatric illnesses, possibly using biomarkers. Here, we investigate longitudinal individual-level predictions of depressive relapse and the level of psychosocial functioning.

**Methods:** Clinical variables (containing detailed symptom profile, previous disease course, as well as environmental and psychological protective and risk factors) and resting-state functional connectivity (rsFC) measures were used to predict relapse and the level of psychosocial functioning after a two-year follow-up interval in 346 patients (240 female) with Major Depressive Disorder (MDD). Random Forest machine learning models were computed to test the incremental predictive capability of clinical and rsFC data compared to a reference model containing confounding variables, and of a multimodal model (combining rsFC and clinical data) compared to the clinical model.

**Results:** Clinical information significantly predicted future psychosocial functioning beyond the reference model (21% versus 12% explained variance, *p* < 0.001). Depression relapse can be predicted by clinical information, however not significantly better than by the reference model alone (64.64% versus 57.70% balanced accuracy, *p* = 0.062). Resting-state data did not yield above-chance accuracies on its own (12% explained variance and 51.72% balanced accuracy) and did not hold incremental predictive value compared to clinical variables for either outcome.

**Conclusions:** Baseline clinical information can be used to predict individual future psychosocial functioning, while rsFC patterns fall short of predicting clinical trajectories in MDD over a two-year interval. Sample size, model complexity and methodological considerations are discussed as potential sources of poor translation of MDD biomarkers.

## Introduction

In psychiatry, there is an urgent need for methods to aid individualized patient care to reach sustained remission. Many patients diagnosed with major depressive disorder (MDD), one of the most prevalent and debilitating psychiatric disorders in adults (GBD 2019 Diseases and Injuries Collaborators, 2022; Gutiérrez-Rojas, Porras-Segovia, Dunne, Andrade-González, & Cervilla, 2020; Santomauro et al., 2021), do not respond sufficiently to current treatments (Fava & Davidson, 1996; Wojnarowski, Firth, Finegan, & Delgadillo, 2019) and relapse is likely (Richards, 2011). For individualized and efficient treatment, it is critical to predict which patients are at risk for a future relapse and need intensive care to reach stable recovery. Currently, this is not possible using demographic and clinical data with traditional group-level inference statistics (Moriarty et al., 2022).

Machine learning (ML) approaches focus on individual-level predictions and on generalizability beyond the analyzed sample by learning complex data patterns (Bzdok & Meyer-Lindenberg, 2018; Hahn, Nierenberg, & Whitfield-Gabrieli, 2017; Janssen, Mourão-Miranda, & Schnack, 2018). Furthermore, ML models hold the advantage of having the capacity to handle a large number of feature variables, enabling the combination of various sources of information. Reflecting the assumed multifactorial relationships of variables involved in psychiatric disorders, it has been repeatedly recommended to implement ML models leveraging both clinical information and neuroimaging-derived biomarkers (Hahn et al., 2017; Iniesta, Stahl, & McGuffin, 2016; Janssen et al., 2018; Tiffin & Paton, 2018). An extensive investigation of various magnetic resonance imaging (MRI) modalities for MDD classification using univariate (Winter et al., 2022) and multivariate methods (Winter et al., 2024) suggests that the individual-level predictive value of these biomarkers is modest in a case-control context. A central question is whether the addition of neuroimaging data brings incremental value to a clinical prognosis, beyond routinely available basic demographic and detailed clinical information. For MDD subjects, Schmaal et al. (2015) did not find that a combination of clinical and functional neuroimaging data reached higher predictive performance than individual domains, while Cearns et al. (2019) found such superiority in combining several modalities including structural neuroimaging, though only in comparison to single biological modalities, and not to clinical data. Hence, current evidence from multimodal prediction studies (Cearns et al., 2019; Schmaal et al., 2015) is conflicting and too sparse to derive recommendations for clinical translation. Further, prior works have not yet covered functional connectivity neuroimaging (Cearns et al., 2019; Schmaal et al., 2015), missed to incorporate between-model incremental tests and used datasets with limited clinical information lacking detailed symptom and risk profiles (Schmaal et al., 2015). Using a qualitatively broad scope of information is important to reflect the multifactorial etiology of MDD.

Resting-state functional connectivity (rsFC) from functional magnetic resonance imaging (fMRI) reflects the default communication pathways of the brain at rest (Sporns, 2012; van den Heuvel & Hulshoff Pol, 2010) and rsFC alterations have been associated with MDD diagnosis (Kaiser, Andrews-Hanna, Wager, & Pizzagalli, 2015; Young et al., 2023) and with future relapse at group level (i.e., using standard inference statistics) (Lythe et al., 2015; Workman et al., 2017). Importantly, rsFC has also emerged as a particularly promising neuroimaging modality to identify cases of MDD on an individual level using ML-based classification (Kambeitz et al., 2017). However, it is currently unclear whether individual rsFC patterns are suited to make predictions about future disease trajectories in MDD and whether predictions can be optimized by combining rsFC data with extensive clinical information spanning broad subdomains of patient characteristics.

A variety of predicted outcome variables has been used across previous studies. Classifications relating to MDD diagnosis status like depressive relapse are most frequently done. Psychosocial functioning is another important operationalization of disease trajectory (Ro & Clark, 2009) as it is regularly impaired in MDD (Fried & Nesse, 2014; Kupferberg, Bicks, & Hasler, 2016; Weightman, Knight, & Baune, 2019) and captures a patient-centered aspect of remission (Kennedy et al., 2016; Zimmerman et al., 2006). Despite several authors recognizing the importance of psychosocial functional recovery (Bakish, 2001; Greer, Kurian, & Trivedi, 2010; Lam, Filteau, & Milev, 2011), it has been largely neglected in treatment effectiveness studies (Kamenov, Cabello, Coenen, & Ayuso-Mateos, 2015).

Building on previous findings (see also Teutenberg et al., 2025), we expect rich clinical variables characterizing severity of symptoms and current disease status (Buckman et al., 2018; Burcusa & Iacono, 2007; Moriarty et al., 2022), prior course of MDD (MacQueen et al., 2000; Moriarty et al., 2022; Richards, 2011), psychological risk and protective factors (Buckman et al., 2018; Burcusa & Iacono, 2007; Cristóbal-Narváez, Haro, & Koyanagi, 2020; Nanni, Uher, & Danese, 2012; Su, Meng, Yang, & D’Arcy, 2022; Watters, Aloe, & Wojciak, 2023), as well as rsFC to be associated with the outcome variables depressive relapse or recurrence in the follow-up period (relapse) and global assessment of functioning (GAF) at follow-up. Thus, we hypothesize the ML algorithm to be able to predict the outcome variables significantly above chance level based on these feature types, both individually and combined and that the accuracy of the combined approach significantly exceeds the prediction accuracy using clinical parameters alone. Furthermore, we consider the importance of implementing sensible model significance tests, accounting for the predictive value of demographic and confounding variables.

## Methods and Materials

### Participants and Design

Data from the longitudinal Marburg/Münster Affective Disorders Cohort Study (MACS, Kircher et al., 2019) were used. The recruitment, data collection procedure and inclusion criteria are described in the Supplements. All subjects provided written informed consent, and the authors assert that all procedures contributing to this work comply with the ethical standards of the relevant national and institutional committees on human experimentation and with the Helsinki Declaration of 1975, as revised in 2008. We included data from 346 participants with a lifetime MDD diagnosis, with available resting-state fMRI scans, basic demographic and clinical characterization at baseline, and available clinical follow-up data approximately two years later (mean interval = 2.18 years, SD = 0.24, range = [1.83;2.99]). Participant characteristics and summary statistics for all control and clinical variables are presented in Table 1 and in the Supplements.

**Table 1.** Sample Characteristics.

| Features | No Relapse<br>(*n* = 175)<br>*M* ± *SD* | Relapse<br>(*n* = 171)<br>*M* ± *SD* | Total<br>(*n* = 346)<br>*M* ± *SD* | *p* |
| --- | --- | --- | --- | --- |
| Site |  |  |  |  |
| Marburg | 83 (47%) | 108 (63%) | 191 (55%) | 0.003* |
| Münster | 92 (53%) | 63 (37%) | 155 (45%) |  |
| Age | 37.93 ± 12.79 | 33.85 ± 12.88 | 35.91 ± 12.98 | 0.003* |
| Sex |  |  |  |  |
| Female | 118 (67%) | 122 (71%) | 240 (69%) | 0.429 |
| Male | 57 (33%) | 49 (29%) | 106 (31%) |  |
| Follow-Up Interval in Years | 2.18 ± 0.21 | 2.19 ± 0.25 | 2.18 ± 0.23 | 0.688 |
| GAF at Baseline (*n* = 341) | 69.47 ± 13.46 | 63.36 ± 12.41 | 66.44 ± 13.29 | <0.001* |
| HAM-D Total Score | 6.70 ± 5.91 | 9.63 ± 6.98 | 8.15 ± 6.62 | <0.001* |
| Antidepressant Medication |  |  |  |  |
| No | 84 (48%) | 57 (33%) | 141 (41%) | 0.006* |
| Yes | 91 (52%) | 114 (67%) | 205 (59%) |  |
| Remission |  |  |  |  |
| Acute | 53 (30%) | 74 (43%) | 127 (37%) | <0.001* |
| Partial Remission | 46 (26%) | 61 (36%) | 107 (31%) |  |
| Full Remission | 76 (43%) | 36 (21%) | 112 (32%) |  |
| Psychiatric Comorbidity |  |  |  |  |
| No | 116 (66%) | 92 (54%) | 208 (60%) | 0.018* |
| Yes | 59 (34%) | 79 (46%) | 138 (40%) |  |
| Number of MDEs (*n* = 332) | 3.01 ± 3.60 | 5.32 ± 9.82 | 4.10 ± 7.32 | 0.006* |
| Recurrence of MDD |  |  |  |  |
| Single MDE | 80 (46%) | 38 (22%) | 118 (34%) | <0.001* |
| Recurrent | 95 (54%) | 133 (78%) | 228 (66%) |  |
| Age of Onset of MDD | 28.32 ± 12.31 | 22.34 ± 10.94 | 25.35 ± 12.01 | <0.001* |
| CTQ Total Score (*n* = 344) | 43.52 ± 15.59 | 45.55 ± 14.25 | 44.53 ± 14.96 | 0.208 |
| LEQ Total Score | 20.50 ± 12.94 | 22.95 ± 19.25 | 21.71 ± 16.39 | 0.167 |
| SSQ Total Score (*n* = 344) | 4.05 ± 0.74 | 3.69 ± 0.83 | 3.87 ± 0.80 | <0.001* |
| PSS Total Score (*n* = 345) | 26.71 ± 9.97 | 31.13 ± 8.77 | 28.90 ± 9.64 | <0.001* |
| RS25 Total Score (*n* = 345) | 118.11 ± 23.89 | 106.49 ± 23.27 | 112.39 ± 24.26 | <0.001* |
| GAF Score at Follow-Up | 79.78 ± 11.93 | 66.42 ± 13.13 | 73.18 ± 14.20 | <0.001* |

*Notes.* All feature variables were recorded at the baseline assessment; Asterisks indicate significant differences between relapse and no relapse groups based on a 5% significance level; GAF = global assessment of functioning, HAM-D = Hamilton Depression Rating Scale, MDE = Major Depressive Episode, MDD = Major Depressive Disorder, CTQ = Childhood Trauma Questionnaire, LEQ = Life Event Questionnaire, SSQ = Social Support Questionnaire, PSS = Perceived Stress Scale, RS25 = Resilience Scale.

### Clinical Characterization and Outcomes

The clinical feature set consisted of 86 variables including information about severity of symptoms (e.g., total scores and item scores of the Beck Depression Inventory [BDI]) and current disease status (e.g., remission status and medication), prior course of MDD (e.g., lifetime number of major depressive episodes (MDEs)), and psychological risk and protective factors, namely childhood adversities, stressful life events, social support, perceived stress, and psychological resilience. No features were entered into the analysis with more than 20% missing data. Imputation was conducted with less than 20% missing, carefully within the cross-validation scheme to avoid data leakage (for details of imputation procedure see Supplements).

A binary relapse outcome variable was defined as “0” for no additional MDE during the interval and “1” for one or more MDEs in the interval. Our dataset did not distinguish between relapse and recurrence, thus the term relapse is used irrespectively of the duration spent in a symptom free state before the recrudescence of depressive symptoms. Psychosocial functioning was represented by the GAF ranging from 1 to 100, which forms Axis V of the multiaxial system of the Diagnostic and Statitical Manual of Mental Disorders DSM-IV-TR (American Psychiatric Association [APA], 2000; Saß, 2003).

### fMRI Data Acquisition and Preprocessing

Two 3T MRI scanners at Marburg and Münster were used to run an 8-minute resting-state sequence, during which subjects were asked to keep their eyes closed and were not presented any stimuli or tasks. A body coil change at the Marburg scanner resulted in a total of three different hardware conditions which were controlled for using dummy variables.

Preprocessing was done using the default volume-based pipeline in the CONN (v18b) toolbox (Whitfield-Gabrieli & Nieto-Castanon, 2012) (http://www.nitrc.org/projects/conn) within SPM12 (https://www.fil.ion.ucl.ac.uk/spm/software/spm12/). Details on scanning parameters, preprocessing and quality check procedures are described in Vogelbacher et al. (2018) and in the Supplements.

We used the Schaefer Atlas with 100 Parcels in 17-Networks (Schaefer et al., 2018), plus 56 subcortical regions (all except for cerebellum and vermis) from the Automatic Anatomical Labeling Atlas 3 (AAL3, Rolls, Huang, Lin, Feng, & Joliot, 2020) for brain parcellation. For each subject, a region of interest (ROI)-to-ROI connectivity matrix of all 156 regions’ timeseries was derived by computing the Fisher transformed bivariate Pearson’s correlations between all regions (Nieto-Castanon, 2020). This resulted in 12090 data points for each subject that were used as the final resting-state functional connectivity features.

### Models and Analysis

Relapse was predicted in a classification approach and GAF in a regression approach, both using a Random Forest as base estimator. Random Forests are ensemble algorithms which allow non-linear functions to be learned while retaining high explainability via their locally linear properties.

For each of the two outcomes, four models were computed for hierarchical comparisons to deduct incremental predictive value of feature modalities (Yarkoni & Westfall, 2017):

1. A reference model was computed in addition to the clinical and neurobiological models to give a basic benchmark for predictions made with general demographic and confounding variables. This model included six variables for relapse (site, two dummy variables for scanner hardware configurations, age and sex of participants, and length of interval between baseline fMRI scans and follow-up interviews) and seven variables for GAF (the six confounders of relapse plus GAF at baseline). For details see Supplements.
2. A clinical model included all variables from the reference model and the 86 clinical parameters, but not the fMRI data, resulting in 92 features. Note that this model has the same number of features in the relapse and GAF versions, as GAF at baseline is present in both clinical models as a clinical feature.
3. A resting-state model included all variables from the reference model and the 12090 data points of fMRI data, but not the clinical parameters, resulting in 12096 features (12097 in the regression).
4. A multimodal model included all variables from the reference model and both the 86 clinical parameters and the 12090 data points of fMRI data, resulting in 12182 features.

The ML analyses were run in PHOTONAI (Leenings et al., 2021) and data preparation, descriptive analyses and visualizations in R (R Core Team, 2023). Using PHOTONAI, a ML-pipeline (Supplements) was built for the cross-validation and model estimation process to ensure that imputation, feature selection and model training were only performed on the training set and subsequently applied to the test data to prevent data leakage. In short, a nested cross-validation with ten inner folds for hyperparameter optimization of maximum tree depth and ten stratified outer folds for model fitting were used. To handle the risk of overfitting, dimensionality reduction (optimized for PCA or feature selection, Supplements) was performed on the rsFC data prior to model training.

To assess significance of differences in performances between models, permutation testing (Combrisson & Jerbi, 2015; Golland & Fischl, 2003) was administered (n=1000 permutations, Supplements). A significance level of p<.05 was defined. Performance differences of clinical to reference, resting-state to reference and multimodal to clinical models were assessed for both outcomes by subtracting permutation distributions of the respective models (Supplements). As predictive capacity was already fairly well established for clinical data (e.g., Dinga et al., 2018; Kessler et al., 2016; J. L. Wang et al., 2014) but largely unclear for rsFC, testing the incremental predictive value of clinical data to rsFC data (and thus a test of the multimodal versus the rsFC model) was not part of our research question.

Permutation feature importance was computed exploratorily in the reference and clinical models as delineated in the Supplements.

Post-hoc simple bivariate associations of the continuous and categorical feature variables in the clinical and reference models with the two outcome variables were tested using standard inference statistics, i.e., Welch’s two sample t-test and Pearson’s chi squared test for relapse and t-test and analysis of variance for GAF.

Hypotheses and analysis plan were preregistered prior to data analysis to reduce researcher degrees of freedom and facilitate replicability of results (Klapwijk, van den Bos, Tamnes, Raschle, & Mills, 2021). The preregistration, all results and a deviation protocol (documenting deviations from the preregistered analysis plan) are publicly available at an online repository (https://osf.io/72gme). Analysis and results description follow the Transparent Reporting of a multivariable prediction model for Individual Prognosis Or Diagnosis (TRIPOD)-Statement (Collins, Reitsma, Altman, & Moons, 2015; Moons et al., 2015).

## Results

### Descriptive Statistics and Bivariate Associations

Descriptive summary statistics for feature variables across relapse groups and GAF scores as a function of feature variables are reported in Table 1 and in the Supplements. In short, relapse rates and GAF scores differed significantly as a function of most clinical features and confounding variables. Relapsing patients also had a significantly lower GAF score at follow-up. Overall, follow-up GAF scores were higher and had a larger variance than the GAF scores at baseline and the no-relapse group had descriptively higher GAF scores at both timepoints (cf. Supplements). There were significant correlations among most clinical variables, as shown in Figure 1.

**Figure 1.**
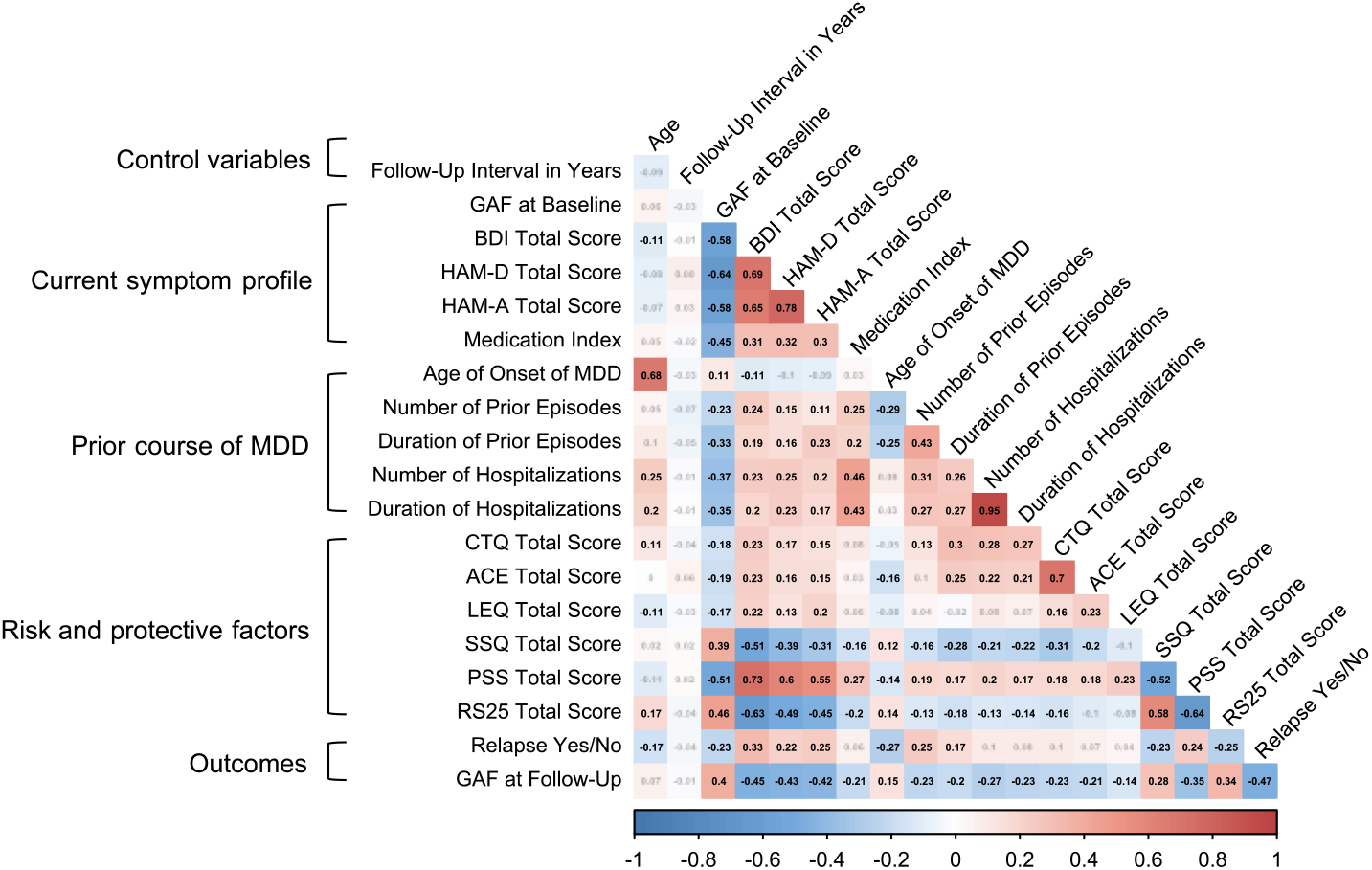
Correlation Matrix of Continuous Features. Note. Matrix cells are color-coded by bivariate Spearman’s correlations. Coefficients that are non-significant on a 5% level are faded out. GAF = global assessment of functioning, BDI = Beck Depression Inventory, HAM-D = Hamilton Depression Rating Scale, HAM-A = Hamilton Anxiety Rating Scale, MDD = Major Depressive Episode, CTQ = Childhood Trauma Questionnaire, ACE = Adverse Childhood Experience questionnaire, LEQ = Life Event Questionnaire, SSQ = Social Support Questionnaire, PSS = Perceived Stress Scale, RS25 = Resilience Scale. The plot was created with Hmisc and corrplot R packages (Harrell Jr, 2023; Wei & Simko, 2021).

### Multivariate pattern prediction of clinical trajectories

Table 2 shows the performance metrics of all eight individual models. Supplementary Figure S1 shows the confusion matrices of all four classification models predicting relapse, giving a detailed depiction of how many cases were correctly and incorrectly classified in both relapse groups. For detailed depictions of the permutation tests complementing the model estimations, see Supplements. Table 3 shows the model comparison parameters.

**Table 2.** Mean Performance Metrics Across All Outer Folds in the Classification of Relapse and Regression of GAF.

| Model | Classification Performance Metrics |  |  | Regression Performance Metrics |  |  |  |
| --- | --- | --- | --- | --- | --- | --- | --- |
| | BAC<br>in % | Sensitivity<br>in % | Specificity<br>in % | MSE | RMSE | MAE | $R^2$ |
| Reference | 57.70<br>(6.62) | 50.82<br>(9.42) | 64.58<br>(13.22) | 176.72<br>(26.18) | 13.26<br>(0.99) | 11.00<br>(1.04) | 0.12<br>(0.12) |
| Clinical | 64.64<br>(10.67) | 65.42<br>(14.14) | 63.86<br>(14.37) | 159.70<br>(26.50) | 12.59<br>(1.08) | 10.38<br>(1.10) | 0.21<br>(0.12) |
| Resting-State | 51.72<br>(5.58) | 50.29<br>(8.77) | 53.14<br>(5.54) | 176.79<br>(26.50) | 13.26<br>(1.00) | 11.00<br>(1.07) | 0.12<br>(0.12) |
| Multimodal | 63.50<br>(9.91) | 63.07<br>(14.55) | 63.92<br>(13.88) | 160.60<br>(27.77) | 12.62<br>(1.15) | 10.40<br>(1.08) | 0.20<br>(0.12) |

*Note.* BAC = balanced accuracy, MSE = mean squared error, RMSE = root mean squared error, MAE = mean absolute error, $R^2$ = explained variance.

**Table 3.** Absolute Differences and Significance of Model Comparisons.

| Comparison | Classification of Relapse |  | Regression of GAF |  |
| --- | --- | --- | --- | --- |
|  | Difference in BAC | *p* | Difference in MSE | *p* |
| Clinical – Reference | 6.94% | 0.062 | -17.02 | <0.001* |
| Resting-State – Reference | 5.98% | 0.882 | 0.07 | 0.562 |
| Multimodal – Clinical | -1.14% | 0.578 | 0.9 | 0.63 |

*Note.* Asterisks indicate significant differences between compared models based on a 5% significance level; corresponding figures are depicted in supplementary Figure S6 and Figure S7; GAF = global assessment of functioning, BAC = balanced accuracy, MSE = mean squared error.

### Reference Model Performance

Basic demographic and confounding variables in the reference models yielded predictions significantly better than chance level both for relapse (balanced accuracy [BAC] = 57.7%, *SD* = 6.62%, *p* = 0.015) and GAF (mean squared error [MSE] = 176.72, *SD* = 26.18, *p* < 0.001). All permutation distributions are shown in the Supplements.

### Clinical Model Performance

The clinical model significantly predicted relapse (BAC = 64.64%, *SD* = 10.67%, *p* < 0.001, Supplements). However, the incremental prediction performance beyond the reference model was not significant (ΔBAC_clinical–reference_ = 6.94%, *p* = 0.062; Table 3 and Supplements).

The clinical model also significantly predicted GAF (MSE = 159.70, SD = 26.50, p < 0.001, Supplements) and explained 21% variance of the GAF follow-up data. In contrast to the relapse predictions, the clinical GAF model did perform significantly better than the GAF reference model (ΔMSE_clinical–reference_ = -17.02, *p* < 0.001, Table 3 and Supplements).

### Resting-State Model Performance

The performance of the relapse resting-state model did not exceed chance level (BAC = 51.72%, *SD* = 5.58%, *p* = 0.278, Supplements). Even on a descriptive level, this model’s ROC curve is close to chance level (Figure 2A). Its performance was descriptively lower than the relapse reference model (ΔBAC_resting-state–reference_ = -5.98%, *p* = 0.882, Table 3 and Supplements).

**Figure 2.**
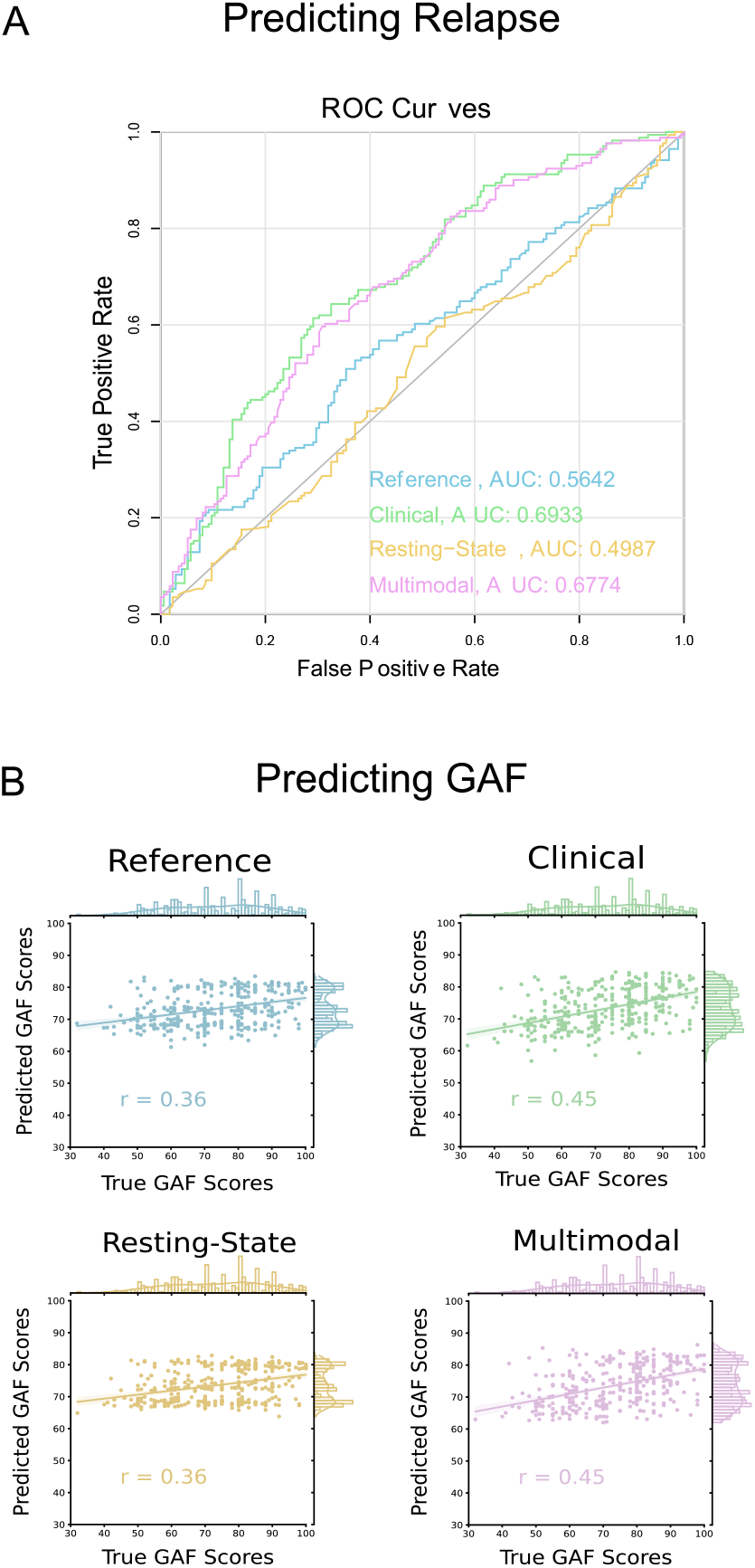
Performance of the Relapse Classification and GAF Regression Models. Note. Panel A shows the Receiver Operating Characteristic (ROC) curves of all four relapse classification models with annotated area under the curve (AUC) values. The grey identity line represents chance level accuracy. The plots were created using cvms and pROC R packages (Olsen & Zachariae, 2023; Robin et al., 2011). Panel B shows the performance of the GAF regression models as scatterplot and least squares lines of the predicted and true GAF scores with marginal histograms and density distributions. The annotated *r* values are the Pearson’s correlation coefficient. The plots were created using seaborn and matplotlib Python packages (Hunter, 2007; Waskom, 2021).

The resting-state model could predict GAF beyond chance level (MSE = 176.79, *SD* = 26.5, *p* < 0.001, Supplements) but its performance did not significantly exceed the GAF reference model (ΔMSE_resting-state–reference_ = 0.07, *p* = 0.562, Table 3 and Supplements).

### Multimodal Model Performance

The multimodal model significantly predicted relapse (BAC = 63.5%, *SD* = 9.91%, *p* < 0.001, Supplements). Descriptively, the relapse clinical and multimodal model performed very similarly to each other, which can be seen in Table 2, the confusion matrices (Supplements) and the ROC curves (Figure 2A). As the difference tests revealed, the multimodal model did not outperform the clinical model (ΔBAC_multimodal–clinical_ = -1.14%, *p* = 0.578, Table 3 and Supplements).

The multimodal model predicted GAF significantly (MSE = 160.60, *SD* = 27.77, *p* < 0.001, Supplements) and explained 20% variance of the GAF follow-up data, but did not outperform the clinical model either (ΔMSE_multimodal–clinical_ = 0.9, *p* = 0.63, Table 3 and Supplements).

Additional performance metrics across all ten outer folds for the prediction of relapse and GAF are summarized in Table 2. In the GAF predictions, it is noteworthy that misestimation in all models is more than 10% of the GAF scale and as Figure 2B reveals, the predicted values fall within a narrower range than the true GAF scores and there are moderate correlations between them.

Both the relapse and GAF clinical models had information from all clinical subdomains as well as single questionnaire items in their top ranked features. Detailed results of the feature importance permutation analysis, the hyperparameter configurations that the algorithm chose, and mean training and test set performance of all models are summarized in the Supplements.

## Discussion

This work investigated multimodal prediction of individual MDD trajectories in an unprecedented scale by integrating functional brain connectivity patterns and comprehensive clinical characteristics. In a large, clinically heterogenous sample, we were able to predict relapse and psychosocial functioning two years into the future using a rich set of clinical data covering detailed symptom profiles, prior disease course and psychological and environmental risk and protective factors. Even when controlling for confounding and basic demographic variables, the predictive performance for psychosocial functioning with the clinical feature set proved to be significant. The rsFC data predicted only psychosocial functioning, but not significantly compared to the confounding variables. Furthermore, rsFC patterns did not add prognostic value to models including only clinical data.

In prior studies, the use of clinical features in multivariate models to predict categorical MDD outcomes generally achieved acceptable to moderate performance (Cearns et al., 2019; Dinga et al., 2018; Schmaal et al., 2015; Teutenberg et al., 2025). Other studies with similar predictions did not benchmark their results from clinical models to a model with confounding variables. Hence their results are not secured against false prognostic value stemming from demographic or other clinically non-relevant information. As our findings indicate, basic demographic and confounding variables already hold substantial predictive value, requiring the need for clinical models to demonstrate additional prognostic capacities. While the hierarchical comparison has been suggested previously (for example by Yarkoni & Westfall, 2017), approaching it with permutation differences to test for significant between-model differences poses an innovative, non-parametric technique to evaluate utility of various feature modalities, though it required careful considerations like the stratified cross-validation. The method used by previous works, i.e., standard permutation testing (Cearns et al., 2019; Dinga et al., 2018; Schmaal et al., 2015), only allows for a significance test of the isolated metric of one model without considering possible confounders or directly comparing models with added feature types.

Our results corroborate Dinga et al. (2018), who also found that a MDD trajectory prediction based on clinical data (i.e., symptom severity) is not improved by the addition of other data types. Also in accordance with the findings at hand, a multimodal model by Cearns et al. (2019) with clinical, sMRI and other biological features was not significantly superior to a clinical model in MDD rehospitalization prediction. Inversely, Schmaal et al. (2015) found that fMRI contrasts from an emotion processing task were nominally better predictors than clinical data and a multimodal model for the prognosis of future chronic course of MDD. Besides their small sample size that bears a risk of overestimation (Flint et al., 2021), chronification as an especially severe outcome may be easier to differentiate from stable remission than relapse of varying severity.

Psychosocial functioning is an important patient-centered outcome in clinical practice that is currently underrepresented in pertinent research (Kamenov et al., 2015; Zimmerman et al., 2006). To the best of our knowledge, this is the first investigation to predict psychosocial functioning of psychiatric patients in a longitudinal design with ML regression, and it was better predictable than binary relapse, even despite controlling for psychosocial functioning at baseline. A continuous measure ranging from excellent functioning over subclinical states to extreme psychosocial disability can more finely resolve individuals’ outcomes and captures a wider variance than dichotomization of the outcome. The fact that related works using dichotomized psychosocial functioning also obtained nominally slightly higher performance than the relapse prediction here potentially indicates that high or low functioning is easier to predict than relapse of clinical magnitude (Koutsouleris et al., 2018; Rodrigues de Aguiar et al., 2023). Further, regression methods allow for the graded assessment of performance with estimations close to or diverging from the true scores accounting for small or large errors. This highlights the benefits of considering continuous or probabilistic outcomes in psychiatric predictive models.

A recent study on MDD trajectory prediction with clinical data suggests that dense feature sets do not improve prediction over a limited set of clinical risk factors (Teutenberg et al., 2025). However, while interpretation of feature importance indices is to be treated with caution because their stability correlates with sample size and complexity (H. Wang, Yang, & Luo, 2016; Yang, Piao, Lai, & Pei, 2017), our results (Supplements) suggest the benefit of including a broad range of information as well as symptom-level data in the analyses. Suicidal ideation and physical symptoms were among the most informative features, which is in line with previous studies (Burcusa & Iacono, 2007) and might point to severity effects on long-term outcomes and the existence of subtypes of MDD with differing risk for unfavorable courses. HAM-A on first rank in the GAF clinical model points to comorbid anxiety being relevant for functional outcomes and replicates longitudinal associations between anxiety symptoms and future depressive symptoms (Jacobson & Newman, 2017).

This work is the first study to utilize multimodal integration of rsFC data in the prediction of individual clinical trajectories in MDD. Furthermore, our sample size is among the largest which have been used for multimodal predictions of MDD trajectories with neuroimaging data (Cearns et al., 2019; Schmaal et al., 2015). In terms of generalizability, our large sample adequately reflects the clinical heterogeneity so that the true utility of our ML model was less likely overestimated than in smaller samples (Flint et al., 2021; Janssen et al., 2018; Schnack & Kahn, 2016).

Conversely, large heterogenous samples necessarily have a more limited possibility to reach high performance (Schnack & Kahn, 2016; Wolfers, Buitelaar, Beckmann, Franke, & Marquand, 2015). Here, different factors may have been important for the prediction of relapse in different clinical subgroups or after short-term versus long-term remission, which may account in part for the mediocre performance. One aspect that still limits representativeness is the culturally homogenous composition of the sample in terms of ethnicity and sociodemographic profiles.

Our analysis was based on broad data characterizing the liability at baseline for future depressive trajectories. However, the nature of our prognostic study did not allow for the inclusion of environmental factors like life events and treatment effects that potentially might have been predictive of the two-year outcomes but occurred after the baseline measurement. Therefore, these kinds of influences could not be addressed in our investigation.

It is likely that despite the automatic feature reduction (Wolfers et al., 2015) of the rsFC data, there were still too many neuronal features that introduced noise into the analysis, causing the observed overfitting (Supplements). This notion is supported by the fact that adding the rsFC information in the multimodal model led to a descriptively poorer performance as compared to the clinical model. The utilized feature reduction poses an intentionally naïve but unbiased and data-driven approach in order to provide the algorithm with as much information as possible (cf. Jollans et al., 2019). Future studies may explore the benefit of theory-driven ROI or combined approaches to reduce the dimensionality to parameters that are most likely mechanistically related to the outcome, performing an expert selection of features (Huys, Maia, & Frank, 2016; Wolfers et al., 2015).

When evaluating clinical utility of single subject predictions, sensitivity and specificity are especially relevant (Arbabshirani, Plis, Sui, & Calhoun, 2017; Wolfers et al., 2015). Here, the sensitivities imply that by adding clinical information to the reference model, more relapse cases tended to be classified correctly, while specificity was not improved (Table 2). The magnitude of the clinical and multimodal relapse prediction models’ metrics (64.64% and 63.50%, though insignificant compared to the reference model) can be interpreted as modest effects (Schnack & Kahn, 2016; for machine learning regression analyses, no effect size guidelines have been proposed to date). Clinical utility can be ascribed to moderately performing predictive models as long as clinicians’ predictions are outperformed (Bzdok & Meyer-Lindenberg, 2018), as found by Koutsouleris et al. (2018). As in many prior studies, no expert predictions were available for our analyses, hence future studies should ask for human prognoses as an important benchmark, wherever possible.

In conclusion, this work contributes to the current debate whether biomarkers can help overcome challenges of precision psychiatry by considering neuroimaging data as input features in ML models. This was the first work to assess individual relapse risk and psychosocial functioning in MDD patients by using a multimodal ML method, including rsFC patterns. In a large, well-curated sample with adequate generalizability, our analyses demonstrated that clinical data is fit to predict psychosocial functioning after two years. Yet, models containing rsFC data fall short of adding meaningful predictive value, rendering a translation into clinical MDD management currently elusive.

## Data Availability

All data produced in the present study are available upon reasonable request to the authors.

