## Supplements for "Predicting Relapse and Psychosocial Functioning in Major Depressive Disorder: A Machine Learning Approach Using Clinical and Resting-State fMRI Data"

### Supplementary Materials

#### Contents

Supplementary Methods

Supplementary Results

References

| | |
|---|---|
| Table S1 | Imputation Rules |
| Table S2 | Summary Statistics for Single Items and Subscales of Questionnaires |
| Table S3 | Summary Statistics of GAF for Categorical Features |
| Table S4 | Top 20 Features Based on Permutation Ranks |
| Table S5 | Best Maximum Depth in Each Fold of Every Model |
| Table S6 | Best Feature Reduction Method in Each Fold of Every Model |
| Figure S1 | Confusion Matrices of the Relapse Classification Models |
| Figure S2 | Development of GAF Scores From Baseline to Follow-Up Assessments of All Participants |
| Figure S3 | Development of GAF Scores From Baseline to Follow-Up Assessments for Relapse and No Relapse Groups |
| Figure S4 | Permutation Distributions of the Relapse Models |
| Figure S5 | Permutation Distributions of the GAF Models |
| Figure S6 | Comparisons of Classification Models for the Prediction of Relapse |
| Figure S7 | Comparisons of Regression Models for the Prediction of GAF |
| Figure S8 | Mean Performance in the Training and Test Sets of the Relapse Models |
| Figure S9 | Mean Performance in the Training and Test Sets of the GAF Models |


### Supplementary Methods

#### Inclusion Criteria

The baseline data collection of this final sample took place between October 2014 and September 2018 and the follow-up phase approximately two years later, between October 2016 and March 2021. General exclusion criteria for study participation were MRI contraindications, non-Caucasian ancestry (due to the initial purpose of the cohorts including genetic investigations), lifetime neurologic diagnosis (including traumatic brain injury, stroke, tumors, dementias, epilepsy, or neuro-inflammation), or lifetime severe somatic illness (including autoimmune, heart, or oncologic diseases, tuberculosis, or hepatitis). For the purpose of the study at hand, we excluded participants with positive or missing answers in the following criteria at the time of baseline fMRI acquisition due to effects on brain functioning: intake of benzodiazepines or z-drugs, stimulants, or substance addiction; and more than 6 months of underweight. In addition, a resting-state quality check was performed as described below, leading to the exclusion of data not meeting quality standards for further analysis. Lastly, participants with questionable compliance (i.e., poor data quality) were excluded as well.

Of 618 eligible depressed subjects that participated in the baseline study, 406 had complete accounts of the outcome variables from the follow-up study. Twenty-two subjects were excluded due to a follow-up interval longer than three years. One additional subject was excluded because of questionable diagnosis. After resting-state quality check, the total sample consisted of 346 participants.

#### Measures of the Control and Feature Variables

Participants were invited for a face-to-face interview with trained personnel at baseline and follow-up to assess their psychiatric history regarding the previous course of their MDD and hospitalizations, current symptoms and patient status, and to fulfill the symptom rating inventories. Afterwards, the study staff decided whether to give a psychiatric diagnosis and rated the GAF score. Paper-pencil questionnaires capturing psychological and clinical data were handed out to participants to be filled in at home and sent back to the departments.

##### Severity of Symptoms and Current Disease Status

All measures in this category were derived from the face-to-face interview at baseline. With the help of the German version of the Structural Clinical Interview for DSM-IV-TR (SCID-IV, Wittchen et al., 1997), the study staff gave the MDD diagnosis and classified the remission status of the patient (full remission, partial remission, or acute), if it is an acute diagnosis or lifetime history of MDD, and whether they suffered from a comorbid psychiatric disorder. In addition, the GAF at baseline (rated as described below) was added as a clinical feature. Additionally, the dataset included information about whether a patient was a psychiatric in-patient at the time of the baseline assessment. Patients were asked to name all psychopharmaceuticals they were taking at the time of the interview and give the respective intake duration and dose; a score indicating medication load was calculated afterwards as follows:

The medication index specifies the level of current psychiatric medication. The assessment of dosage of acute medication was done during the interviews at the baseline study point. Afterwards, the reported medication was categorized according to the active ingredients (e.g., selective serotonin reuptake inhibitors or benzodiazepines; and somatic drugs were categorized as contraceptives, thyroid, or diabetes medication). Established dosage-dependent cutoffs (Benkert & Hippius, 2021; Reynolds, 2008) were then used to code all active ingredients of the psychiatric medication as 0, 1, or 2. Finally, the scores were summed up, composing the final medication index. This procedure has been used numerous times in previous publications (e.g., Goltermann et al., 2022; Redlich et al., 2014; Redlich et al., 2015).

In addition, one variable indicated whether the patient was taking an antidepressant at baseline. From the same session, the total scores and all individual items of the Structured Interview Guide for the Hamilton Depression Rating Scale with Atypical Depression Supplement (SIGH-ADS, Williams & Terman, 2003) and the Beck Depression Inventory (BDI, Beck et al., 1961; Hautzinger et al., 1994) were used as features for a more fine-grained measure of depressive symptoms. In addition, research findings point to the assumption that certain depressive symptoms like sleep disturbances and suicidality are especially predictive of MDD relapse (Burcusa & Iacono, 2007). Further, the item score sum of the German translation of the Hamilton Anxiety Rating Scale (HAM-A, Hamilton, 1959; Strauß, 2005) was used as a feature. For this rating and all following questionnaires, no individual items were added as features because the probable disadvantages of computational load, multicollinearity, and increase in dimensionality outweighed the estimated use of additional information from single items.

##### Course of Depressive Disorder

These features were all obtained from the baseline interview and included the following: retrospective reports from the participant about age of onset of depressive symptoms, lifetime number and cumulative duration of MDEs and hospitalization (including all in-patient and day-care admissions due to their MDD), and whether the MDD was classified as recurrent by the study staff.

#### Risk and Protective Factors

All measures were part of the self-report questionnaire battery. Childhood adversities were assessed with the German version of the Adverse Childhood Experiences questionnaire, accounting for one feature (ACE, Felitti et al., 1998; Wingenfeld et al., 2011), and the German version of the Childhood Trauma Questionnaire (CTQ, Bernstein et al., 2003; Klinitzke et al., 2012) with total and subscores accounting for eight features. In addition, the negative, positive, and overall total events scores from the German translation of the Life Event Questionnaire (LEQ, Norbeck, 1984) were included as three features in the analysis. The sums of the German social support questionnaire „Fragebogen zur sozialen Unterstützung" (SSQ, Fydrich et al., 1987) and the German version of the Perceived Stress Scale (PSS, Cohen et al., 1983; Klein et al., 2016), as well as the subscale sums and the total sum of the German version of the Resilience Scale (RS25, Wagnild & Young, 1993; Leppert et al., 2008) were also added to the feature set.

#### Confounding Variables in the Reference Models

The confounding variables in the Reference model included the site of data collection (Marburg or Münster), and the three different hardware configurations, because of possible effects on resting-state functional imaging despite the quality assurance protocol (Yu et al., 2018). This resulted in two variables dummy-coding for the three setups described above that were included in the analysis. Age and sex of participants are also known to influence resting-state data due to effects on lateralization (Agcaoglu et al., 2015) and complexity (Dong et al., 2018) of functional signals and may also correlate with relapse risk (Kessing, 1998), thus they were controlled for in the Reference model as well. Because with longer follow-up duration, cumulative relapse rates are higher (Solomon et al., 2000), length of interval between baseline fMRI session and follow-up interview in years was also included here. Lastly, GAF score at baseline was also a confounder in the Reference model predicting GAF score at follow-up to account for influences on the follow-up value.

### Measures of the Outcome Variables

#### Depressive Relapse

The diagnostic interview was repeated at follow-up, extended by a part about the participant's depressive symptoms in the interscan interval, conducted with the help of a life chart method (Post et al., 1988). This part of the interview asked about past events, mental well-being, treatments, and impairment since the baseline assessment and the interviewer rated the severity of the disorder over time on a scale from -3 (severe depression) to +3 (severe mania), giving an overview of asymptomatic, subsyndromal, dysthymic, depressive, hypomanic, or manic phases. Afterwards, the experimenter rated whether a relapse had taken place according to the criteria of the DSM-IV using the SCID-IV (Wittchen et al. 1997), which require at least two months of remission before the reoccurrence of depressive symptoms. The binary variable was defined as "0" for no additional MDE during the interval and "1" for one or more MDEs in the interval. Seventeen patients who were chronically depressed throughout the entire follow-up period were also coded as "1", since they were not free of depression, although in these cases no new MDE could be diagnosed. Seven patients with a dysthymic course were rated as "0" because the severity of symptoms did not reach MDD criteria.

#### Psychosocial Functioning

During the follow-up interview, the experimenter also gave a rating of the GAF ranging from 1 to 100, which forms Axis V of the multiaxial system of the DSM-IV-TR (American Psychiatric Association [APA], 2000; Saß, 2003). The realms occupational, psychological, and social functioning are considered for this rating. It follows the hypothesis of a continuum from mental health to mental illness, with 1 being the lowest and 100 the highest possible level of functioning. The 100-point scale is divided into 10 intervals that each contain a description of symptoms and functioning for the rater to categorize the patient's condition. The exact value within the decile allows a finely graded assessment.

#### fMRI Data Acquisition and Preprocessing

The fMRI-acquisition procedure and quality assurance protocol is described in detail in Vogelbacher et al. (2018). Participants did functional resting-state MRI measures at the end of a 45-minute scanning procedure including also sMRI, DTI, and task-based fMRI. During the 8-minute resting-state measurement, subjects were asked to keep their eyes closed and were not presented any stimuli or tasks. The devices were Tim Trio (Marburg) and Prisma (Münster) 3T whole body MRI scanners manufactured by Siemens Healthcare situated in Erlangen, Germany. In 2016, the body coil in Marburg was changed, resulting in three different hardware configurations for this sample: Marburg with old body coil, Marburg with new body coil, or Münster. In Münster, the scanner's prescan normalize option was activated in all cases of this sample to correct for non-uniform receiver coil profiles prior to imaging.

A T2*-weighted echo planar imaging (EPI) sequence was used for functional imaging, acquiring 237 interleaved and ascending measurements that were tilted -20° against the anterior and posterior commission alignment. The acquisition settings comprised a base resolution of 64, a bandwidth of 2232 Hz/px, an echo spacing of 0.51 ms, an EPI factor of 64, a BOLD threshold of 4, a repetition time (TR) of 2000 ms, and an echo time (TE) of 30 ms (Marburg) and 29 ms (Münster). The parameters for the 33 slices covering the whole brain were a slice thickness of 3.8 mm, a field of view (FOV) of 210 mm, a spatial resolution of 3.3 x 3.3 x 3.8 mm3, and a flip angle of 90°.

For the preprocessing of the fMRI data, the default pipeline for volume-based analyses in the CONN (v18b) MATLAB toolbox was followed (Whitfield-Gabrieli & Nieto-Castanon, 2012) (http://www.nitrc.org/projects/conn). This pipeline involved several steps. First, the realign and unwarp procedure from SPM12 (Andersson et al., 2001) was used to co-register and resample all scans to the reference image. This step also accounted for potential susceptibility distortion-by-motion interactions and field inhomogeneity within the scanner (Nieto-Castanon, 2020). Next, the SPM12 slice-timing correction (Henson et al., 1999) was applied to correct temporal misalignment between the slices via time-shifting and resampling the functional data to match the middle of each acquisition time (TA). Subject motion in the scanner exceeding a framewise displacement of 0.5 mm or observed global BOLD signal changes above three standard deviations were used to identify potential outliers (conservative settings, Nieto-Castanon, 2020). The third step comprised the unified segmentation and normalization procedure from SPM12 (Ashburner & Friston, 2005) during which the data were normalized into standard Montreal Neurological Institute (MNI) space and segmented in grey and white matter and cerebrospinal fluid (CSF) tissue classes (Nieto-Castanon, 2020). Lastly, to increase the BOLD signal-to-noise-ratio and reduce the influence of residual variability, the data was smoothed via spatial convolution with a Gaussian kernel of 8 mm full width at half maximum (FWHM, Nieto-Castanon, 2020).

The next part of the pipeline was the denoising of the fMRI data. The default anatomical component-based noise correction (aCompCor) from CONN (Nieto-Castanon, 2020) linearly regressed out five principal components for noise from white matter and cerebrospinal fluid areas, 12 parameters for motion of the subject (three translation and three rotation parameters plus their three respective first order derivates), as well as potential confounding effects of outlier scans. To control for further noise sources from head motion, physiological or other influences, the last step was temporal band-pass filtering of the BOLD signal, keeping 0.008 Hz – 0.09 Hz frequencies.

The resting-state data underwent a final quality check. For each subject, individual voxel-to-voxel connectivity distribution histograms were checked for visible irregularities in the form of skewed distributions indicating artificial hyperconnectivity. No irregularities were found in this process. Subjects with less than 150 non-scrubbed scans did not reach a minimum of five minutes valid resting-state and were excluded from the analysis.

#### Random Forest Algorithm

PHOTONAI used the default parameters of the RandomForestClassifier and RandomForestRegression algorithms specified in scikit-learn (Pedregosa et al., 2011), unless otherwise indicated.

First, for imputation of the data, the SimpleImputer from scikit-learn was applied to all models with missing values being imputed with the mean of the respective feature of the training set (SimpleImputer, strategy = mean). The four models with brain data then branched the resting-state features off for a dimensionality reduction process due to the high number of these features to counteract overfitting. As generalizability beyond the sample at hand is the goal of predictive modeling, it is of paramount importance to control for overfitting as to not fit the model perfectly to the sample data while losing all generalizability to unseen data. Complex models with high-dimensional data easily overfit and thus, dimensionality needs to be reduced and complexity regularized or penalized (Huys et al., 2016). A selection amongst all possible resting-state connections also makes sense in the mechanistic view of trajectory prediction, as it is not reasonable to assume that all connectivity features carry information about depressive course progression, but rather only a fraction of mechanistically relevant ones.

This process implemented a Switch element to optimize for the reduction method which was either principal component analysis (PCA) or feature selection. PCA decomposes a multivariate dataset in orthogonal components to explain the maximum possible amount of variance. All components were kept with the parameter settings. Feature selection with the SelectPercentile component only keeps a certain percentile of the features with the highest univariate association with the target variable. In our analysis, these were the top 5% or top 15% of features. The test statistics of this function are ANOVA F Score for classification (f_classif) and F Score for regression (f_regress). The Switch means that all three reduction methods were tested and then the optimal one was chosen to proceed with. On the second branch, all confounders and in the Multimodal model also all clinical variables were kept without dimensionality reduction due to the smaller number and better signal-to-noise ratio of these features. For the training process, the two branches were recombined, feeding all clinical and control variables and the subset of best resting-state features to the algorithm. In the Reference and Clinical models, no dimensionality reduction was performed, therefore the second branch of the Resting-State and Multimodal models resembles these feature sets to permit better comparability between the different models.

The Random Forest model with the base estimator DecisionTreeClassifier or DecisionTreeRegressor was fit in a ten-fold nested cross-validation procedure (Kohavi, 1995; Varma & Simon, 2006). This means that for the training process, the dataset was divided into ten stratified folds for cross-validation, so that 90% were used for training the model and 10% for testing it, repeated ten times until all subsets were used as test data once. In order to have the same cases in the ten folds of each of the four classification models and similar distributions of Relapse and No Relapse cases across all folds, the folds were stratified according to the Relapse variable with a fixed random state. The same was done for the four regression models, however instead of classic stratification, which is not applicable to continuous outcome variables, sorted stratification according to GAF was implemented. Another level of cross-validation folds was added for hyperparameter optimization of the algorithm to conduct nested cross-validation: Before estimating the models in the outer folds, each of the ten outer folds was further divided into ten inner folds with a test set size of 10%. Applying nested cross-validation for hyperparameter optimization is necessary to prevent data leakage and misestimation (Cearns et al., 2019; Varma & Simon, 2006). The hyperparameter to optimize for was the maximum depth of the tree with possible values [2, 4, 6, 8, 10, 20]. Grid search was implemented to optimize the hyperparameter, which means that all six configurations were tested. The remaining specified hyperparameters had fixed values as follows. In each fold of the cross-validation, 100 trees were calculated. The minimal number of samples to allow a split and the minimum of samples in a leaf were set to two and one, respectively, meaning that splits were performed until either all nodes were pure or only one case was left in an end node (leaf), as long as the maximum depth was not exceeded; these are the most unrestricted settings. The number of samples to draw for each bootstrap sample to build each individual tree was set to the sample size, which is the default in scikit learn. This means that each tree was created from a new random sample with the size of 311 or 312 (346 × 90%), where some cases were present multiple times and others not at all due to bootstrapping. The size of the subset of features to be considered at each split was not restricted (max_features = None), meaning all features that remained after recombining the above-mentioned branches were kept in the analysis. While it created less variation between the trees than intended in the original Random Forest method, we decided on this procedure to present every tree the same clinical and control information, plus the most promising selected resting-state data in the respective models. In addition, in scikit-learn's implementation, "[the] features are always randomly permuted at each split. Therefore, the best found split may vary […]", even between the trees of the same fold when the subset of features is not restricted (https://scikit-learn.org/stable/modules/generated/sklearn.ensemble.RandomForestClassifier.html.) After the inner cross-validation found the best hyperparameter configuration, the 100 trees of the respective outer cross-validation training set were generated to fit the model. The method used by the algorithm to fit the model and search for the best tree splits was Gini impurity in the classification models and MSE in the regression models. The metric used for optimization was BAC for the classification models and MSE for the regression models. Instead of the majority vote for classification from the original publication (Breiman, 2001), to aggregate over all 100 trees of each training data set, scikit-learn's implementation uses the average of all trees' probabilistic predictions based on leaf class distribution. The probability threshold to decide for class 0 (No Relapse) or 1 (Relapse) was 0.50.

This whole procedure was repeated independently with all ten outer folds. The final performance metrics displayed in Table 2 and Table 3 are the averages over all folds.

#### Permutation Testing of Between-Model Performance Differences

For each model, 1000 permutations with shuffled labels were run. To compare the Relapse Clinical model to the Relapse Reference model, the BAC of every permuted Reference model was subtracted from the corresponding BAC of the Clinical model, generating a null distribution of performance differences. This was feasible due to the stratified folds in which the same cases are in the same folds of all different models. The exact p-value of the observed difference between the original Clinical and Reference models' BAC in this distribution was evaluated on a significance level of 5%. This procedure was also performed to compare the Resting-State model to the Reference model and the Multimodal model to the Clinical model. To compare performances between the models in the regression scenario, the same comparisons were done for the MSE metric.

#### Feature Importance

To calculate feature importance in the Reference and Clinical models, the models were re-run 100 times for each feature. The respective feature's values were randomly shuffled to dissolve any correlation with the outcome variable. The normalized mean drop in performance when one features was eliminated compared to the actual model performance was used to create a ranking of the features' importances. Due to a very large computational load, this procedure was refrained from for the four models containing rsFC features. This permutation importance method was chosen because the decrease in impurity method is biased in favor of continuous variables (Nembrini et al., 2018).

### Supplementary Results

#### Descriptive Results

No feature had more than 12% missing data and no subject had more than six missing feature values. Table S2 summarizes means and standard deviations of all single items and questionnaire subscales that composed part of the clinical features. Table S3 shows means and standard deviations of GAF as a function of the levels of the categorical clinical and confounding features. There were significant differences in GAF score with regard to lifetime diagnosis, remission, and inpatient treatment status, comorbidity, antidepressant intake and recurrent MDD diagnosis at baseline (Table S3). Figures S2 and S3 show the development of GAF scores for all participants and for relapse and no-relapse groups, respectively.

#### Multivariate pattern prediction of clinical trajectories

The permutation runs that complemented the model estimations resulted in a permutation distribution for each of the eight models, as shown in Figures S4 and S5. Only 994 instead of 1000 permutations could be generated in the multimodal relapse model due to intractable runtime errors. Consequently, the comparison of the multimodal relapse model to the clinical relapse model using a permutation difference approach (Figure S6) is also based on 994 permutations instead of 1000.

#### Feature Importance

Feature importance was only calculated for the Reference and Clinical models of Relapse and GAF prediction due to the large number of features in the Resting-State and Multimodal analyses causing high computational costs. Table S4 shows the top 20 ranks of permutation importance. The Reference models ranked age on first (Relapse) and second (GAF) rank with GAF at baseline being on first rank in the GAF Reference model. GAF at baseline still has a relatively high permutation importance in the Clinical GAF model (rank 17). Apart from this, in both Clinical models, no control variables were present among the top 20 features. Instead, variables reflecting symptom severity (including single items), prior course of MDD, and all domains of risk and protective factors (childhood maltreatment, recent life events, perceived stress, social support, and psychological resilience) are present in the list.

#### Hyperparameter Configurations

Analysis of the hyperparameter configurations that the algorithm chose (Table S5 for maximum depth and Table S6 for feature reduction methods) reveals that in all folds of the four Reference and Clinical models, a shallow depth between two and eight was chosen, whereas in the analyses with rsFC data, maximum depth of 20 occurred several times. In all folds of the Resting-State and Multimodal GAF prediction models, PCA was chosen for feature reduction and in the Relapse prediction models with rsFC data, the percentile method was chosen often in the Resting-State model folds and once in the Multimodal model. PCA reduces the rsFC features the most, arguing for a preference of the most restricting feature selection method, possibly reflecting a floor effect.

#### Training and Test Set Performance

Figures S8 and S9 show the mean training and test set performance of all models. The largest discrepancies between training and test set performance are apparent in the relapse resting-state model with more than 40% difference in balanced accuracy, and in the GAF multimodal model with more than 50% difference in mean squared error. A much higher performance in the training than in the test set is a sing for overfitting.

### Table S1

*Imputation Rules*

| Measure | Imputation rule |
|---|---|
| *Severity of Symptoms and Current Disease Status* | |
| HAM-D | Imputation with similar record of up to 2 missing items |
| BDI | Imputation with similar record of up to 2 missing items |
| HAM-A | Imputation with similar record of 1 missing item |
| *Risk and protective factors* | |
| CTQ | No imputation after subscale imputation |
| CTQ subscales | Imputation with similar record of 1 missing item |
| ACE | No imputation |
| LEQ | No imputation |
| SSQ | Imputation with similar record up of to 4 missing items |
| PSS | Imputation with similar record of up to 3 missing items |
| RS25 | No imputation after subscale imputation |
| RS25 subscales | Imputation with similar record of up to 2 missing items in Personal Competence scale and up to 3 missing items in Acceptance scale |

*Note*. HAM-D = Hamilton Depression Rating Scale, BDI = Beck Depression Inventory, HAM-A = Hamilton Anxiety Rating Scale, CTQ = Childhood Trauma Questionnaire, ACE = Adverse Childhood Experience questionnaire, LEQ = Life Event Questionnaire, SSQ = Social Support Questionnaire, PSS = Perceived Stress Scale, RS25 = Resilience Scale.

### Table S2

*Summary Statistics for Single Items and Subscales of Questionnaires*

| Variable | *n* | Mean (*SD*) | Variable | *n* | Mean (*SD*) |
|---|---|---|---|---|---|
| BDI Item 1 | 346 | 0.76 (0.81) | HAM-D Item a4 | 346 | 0.21 (0.52) |
| BDI Item 2 | 346 | 0.72 (0.95) | HAM-D Item a5 | 346 | 0.33 (0.61) |
| BDI Item 3 | 346 | 0.80 (0.86) | HAM-D Item 6 | 346 | 0.47 (0.76) |
| BDI Item 4 | 345 | 1.00 (0.85) | HAM-D Item 7 | 346 | 0.51 (0.68) |
| BDI Item 5 | 346 | 0.76 (0.73) | HAM-D Item 8 | 346 | 0.33 (0.67) |
| BDI Item 6 | 346 | 0.67 (0.99) | HAM-D Item a6 | 346 | 0.26 (0.70) |
| BDI Item 7 | 345 | 0.79 (0.79) | HAM-D Item 9_SF | 346 | 0.51 (0.57) |
| BDI Item 8 | 346 | 1.00 (0.84) | HAM-D Item 9 | 346 | 0.80 (0.95) |
| BDI Item 9 | 346 | 0.46 (0.60) | HAM-D Item a7 | 346 | 1.10 (1.30) |
| BDI Item 10 | 345 | 0.76 (0.95) | HAM-D Item 10 | 346 | 0.55 (0.80) |
| BDI Item 11 | 345 | 0.84 (0.91) | HAM-D Item 11 | 346 | 0.29 (0.67) |
| BDI Item 12 | 345 | 0.64 (0.78) | HAM-D Item 12 | 346 | 0.70 (0.91) |
| BDI Item 13 | 345 | 1.10 (0.93) | HAM-D Item 13 | 346 | 0.73 (0.97) |
| BDI Item 14 | 346 | 0.94 (1.00) | HAM-D Item 14 | 346 | 0.22 (0.48) |
| BDI Item 15 | 345 | 1.10 (0.83) | HAM-D Item 15 | 346 | 0.01 (0.14) |
| BDI Item 16 | 344 | 1.00 (0.87) | HAM-D Item 16 | 346 | 0.27 (0.53) |
| BDI Item 17 | 346 | 1.00 (0.80) | HAM-D Item 17 | 346 | 0.20 (0.58) |
| BDI Item 18 | 346 | 0.35 (0.66) | HAM-D Item 18 | 346 | 0.52 (0.76) |
| BDI Item 19 | 345 | 0.31 (0.75) | HAM-D Item a8 | 346 | 0.26 (0.66) |
| BDI Item diet | 345 | 0.24 (0.43) | HAM-D Item 19 | 346 | 0.19 (0.59) |
| BDI Item 20 | 346 | 0.46 (0.60) | HAM-D Item 20 | 346 | 0.05 (0.26) |
| BDI Item 21 | 345 | 0.91 (1.00) | HAM-D Item 21 | 346 | 0.09 (0.33) |
| HAM-D Item 1 | 346 | 0.99 (1.10) | CTQ Emotional Abuse | 346 | 10.82 (5.03) |
| HAM-D Item 2 | 346 | 0.73 (1.00) | CTQ Physical Abuse | 346 | 6.48 (2.80) |
| HAM-D Item a1 | 346 | 0.60 (1.00) | CTQ Sexual Abuse | 345 | 6.48 (3.48) |
| HAM-D Item 3 | 346 | 0.55 (0.79) | CTQ Emotional Neglect | 345 | 13.08 (5.17) |
| HAM-D Item 4 | 346 | 0.18 (0.40) | CTQ Physical Neglect | 346 | 7.71 (2.98) |
| HAM-D Item 5a | 329 | 0.02 (0.17) | LEQ Positive Events | 346 | 9.49 (8.73) |
| HAM-D Item 5b | 7 | 1.70 (0.49) | LEQ Negative Events | 346 | 12.23 (12.31) |
| HAM-D Item a2 | 346 | 0.09 (0.32) | RS25 Acceptance | 345 | 32.14 (8.98) |
| HAM-D Item a3 | 346 | 0.23 (0.57) | RS25 Personal Competence | 345 | 80.28 (17.11) |

*Notes*. HAM-D Item 5 was either answered on Item 5a or 5b. BDI = Beck Depression Inventory, HAM-D = Hamilton Depression Rating Scale, CTQ = Childhood Trauma Questionnaire, LEQ = Life Event Questionnaire, RS25 = Resilience Scale.

#### Table S3

*Summary Statistics of GAF for Categorical Features*

| Features | GAF Mean (*SD*) | *p* |
|---|---|---|
| *Control variables* | | |
| Site | | |
| Marburg | 73.61 (15.23) | |
| Münster | 72.65 (12.83) | 0.524 |
| MRI Device | | |
| Marburg old | 74.63 (15.34) | |
| Marburg new | 69.88 (14.41) | |
| Münster | 72.65 (12.83) | 0.136 |
| Sex | | |
| male | 73.93 (14.58) | |
| female | 72.84 (14.04) | 0.517 |
| *Severity of Symptoms and Current Disease Status* | | |
| Antidepressant Medication | | |
| No | 76.82 (13.80) | |
| Yes | 70.67 (13.95) | <0.001 |
| Remission | | |
| Acute | 67.35 (13.65) | |
| Partial Remission | 72.46 (13.03) | |
| Full Remission | 80.46 (12.64) | <0.001 |
| Lifetime or Acute Diagnosis | | |
| History of MDD Diagnosis | 69.58 (14.07) | |
| Acute MDD Diagnosis | 77.82 (12.99) | <0.001 |
| Psychiatric Comorbidity | | |
| No | 75.90 (13.52) | |
| Yes | 69.07 (14.25) | <0.001 |
| In-Patient Treatment (*n* = 344) | | |
| No | 74.28 (14.01) | |
| Yes | 69.26 (14.15) | 0.01 |
| *Course of the Depressive Disease* | | |
| Recurrence of MDD | | |
| Single MDE | 76.76 (14.49) | |
| Recurrent | 71.32 (13.71) | 0.001 |

*Notes*. All feature variables were recorded at the baseline assessment. GAF = global assessment of functioning at follow-up.

#### Table S4

*Top 20 Features Based on Permutation Ranks*

| Rank | Relapse Reference | Relapse Clinical | GAF Reference | GAF Clinical |
|---|---|---|---|---|
| 1 | Age | BDI Total | GAF Score | HAM-A Total |
| 2 | Site | DepEp | Age | BDI Total |
| 3 | Dummy Bodycoil 1 | PSS | Interval | HAM-D Total |
| 4 | Sex | BDI Item 9 | Dummy Bodycoil 1 | HAM-D Item 9 SF |
| 5 | Dummy Bodycoil 2 | RS25 Personal Competence | Site | HAM-D Item 9 |
| 6 | Interval | HAM-D Item 6 | Dummy Bodycoil 2 | HAM-D Item a7 |
| 7 | | DurHosp | Sex | BDI Item 21 |
| 8 | | SSQ | | DurHosp |
| 9 | | LEQ Negative Events Score | | DepEp |
| 10 | | BDI Item 3 | | CTQ Physical Neglect |
| 11 | | BDI Item 17 | | CTQ Total |
| 12 | | CTQ Physical Neglect | | HAM-D Item 10 |
| 13 | | RS25 Total | | Hosp |
| 14 | | CTQ Emotional Neglect | | HAM-D Item 1 |
| 15 | | CTQ Total | | RS25 Personal Competence |
| 16 | | BDI Item 7 | | CTQ Emotional Neglect |
| 17 | | LEQ Total Events Score | | GAF Score |
| 18 | | BDI Item 6 | | SSQ |
| 19 | | HAM-D Item 9 | | LEQ Negative Events Score |
| 20 | | BDI Item 4 | | HAM-D Item 2 |

*Note.* Ranks were calculated by normalized mean drop in balanced accuracy. GAF = global assessment of functioning, BDI = Beck Depression Inventory, CTQ = Childhood Trauma Questionnaire, DepEp = number of prior depressive episodes, DurHosp = cumulative duration of prior hospitalizations, HAM-A = Hamilton Anxiety Rating Scale, HAM-D = Hamilton Depression Rating Scale, Hosp = number of prior hospitalizations, Interval = follow-up interval in years, LEQ = Life Event Questionnaire, PSS = Perceived Stress Scale, RS25 = Resilience Scale, Site = Marburg or Münster, SSQ = Social Support Questionnaire.

#### Table S5

*Best Maximum Depth in Each Fold of Every Model*

| | Relapse | | | | GAF | | | |
|---|---|---|---|---|---|---|---|---|
| Fold | Ref | Clin | RS | Multi | Ref | Clin | RS | Multi |
| 1 | 2 | 2 | 2 | 2 | 2 | 8 | 2 | 2 |
| 2 | 2 | 8 | 20 | 2 | 2 | 2 | 2 | 8 |
| 3 | 2 | 2 | 20 | 10 | 2 | 2 | 2 | 6 |
| 4 | 4 | 2 | 4 | 10 | 2 | 4 | 2 | 20 |
| 5 | 4 | 4 | 2 | 20 | 2 | 4 | 10 | 4 |
| 6 | 2 | 2 | 2 | 8 | 2 | 4 | 2 | 6 |
| 7 | 4 | 2 | 20 | 2 | 2 | 2 | 2 | 10 |
| 8 | 4 | 2 | 20 | 2 | 2 | 4 | 2 | 4 |
| 9 | 6 | 2 | 8 | 10 | 2 | 2 | 2 | 6 |
| 10 | 4 | 2 | 20 | 2 | 2 | 2 | 2 | 2 |

*Note.* GAF = global assessment of functioning, Ref = Reference, Clin = Clinical, RS = Resting-State, Multi = Multimodal.

#### Table S6

*Best Feature Reduction Method in Each Fold of Every Model*

| | Relapse | | GAF | |
|---|---|---|---|---|
| Fold | Resting-State | Multimodal | Resting-State | Multimodal |
| 1 | 5% | PCA | PCA | PCA |
| 2 | 15% | PCA | PCA | PCA |
| 3 | 15% | PCA | PCA | PCA |
| 4 | 15% | 15% | PCA | PCA |
| 5 | 5% | PCA | PCA | PCA |
| 6 | 5% | PCA | PCA | PCA |
| 7 | PCA | PCA | PCA | PCA |
| 8 | 5% | PCA | PCA | PCA |
| 9 | PCA | PCA | PCA | PCA |
| 10 | 5% | PCA | PCA | PCA |

*Note.* GAF = global assessment of functioning, Ref = Reference, Clin = Clinical, RS = Resting-State, Multi = Multimodal, 5%/15% = feature selection according to univariate association, PCA = principal component analysis.

#### Figure S1

*Confusion Matrices of the Relapse Classification Models*

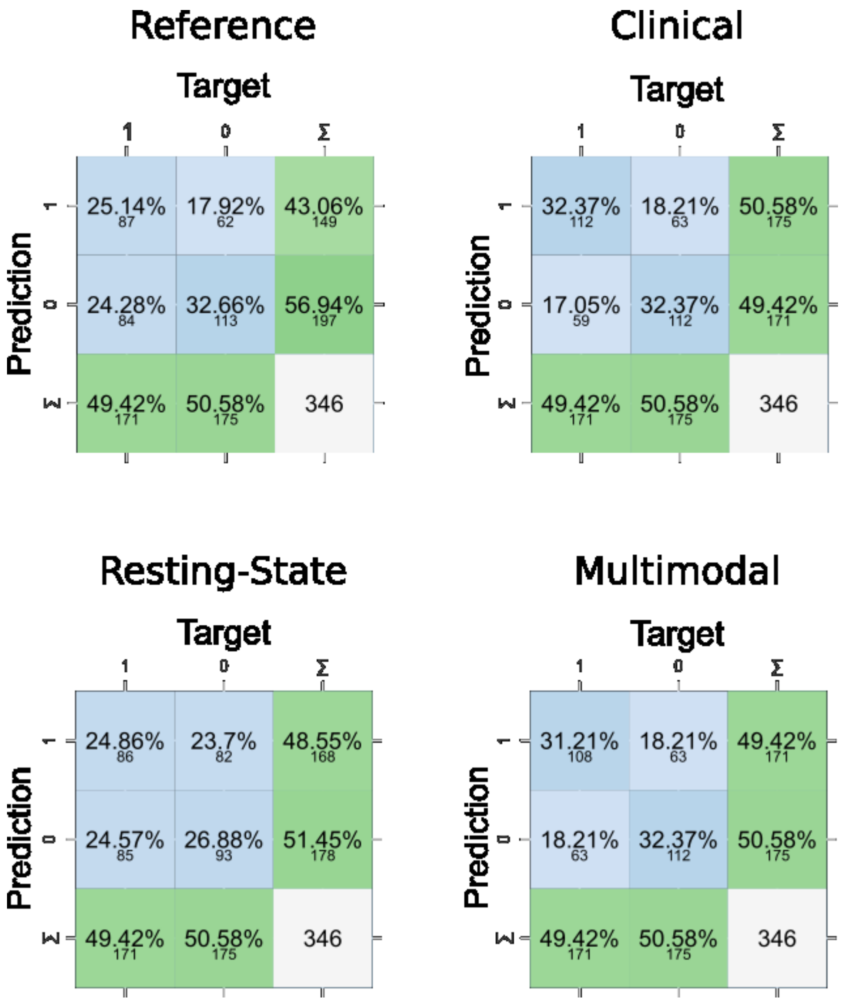


*Note.* Blue panels indicate the number of correctly and incorrectly classified cases. Green panels show the marginal totals. 0 = no relapse, 1 = relapse, ∑ = totals. Below the percentages, absolute values are displayed in each cell.

### Figure S2

*Development of GAF Scores From Baseline to Follow-Up Assessments of All Participants*

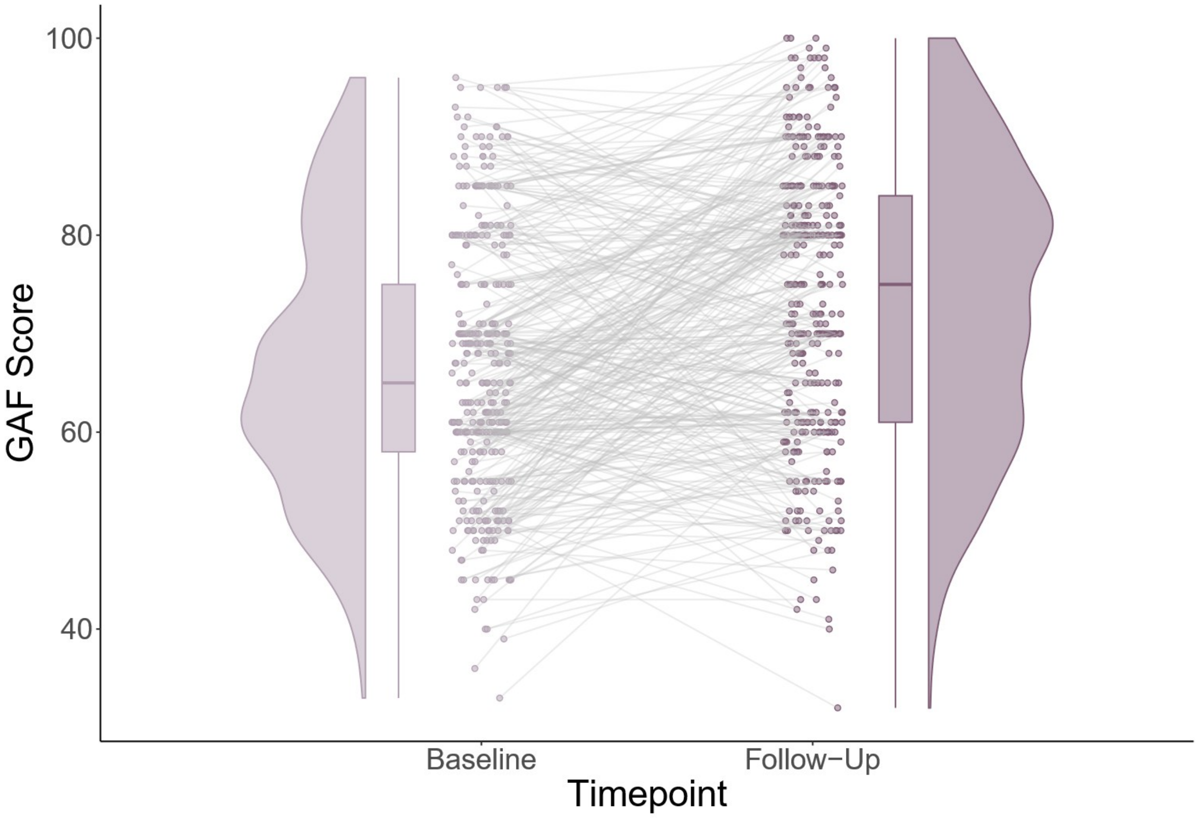


*Note.* Points refer to individual scores, with grey lines connecting the two values of one person. For each timepoint, vertical boxplots and density distributions were added. Five participants were excluded from this figure due to missing values in GAF score at baseline. The plot was created with the raincloudplot R package (van Langen, 2023).

#### Figure S3

*Development of GAF Scores From Baseline to Follow-Up Assessments for Relapse and No Relapse Groups*

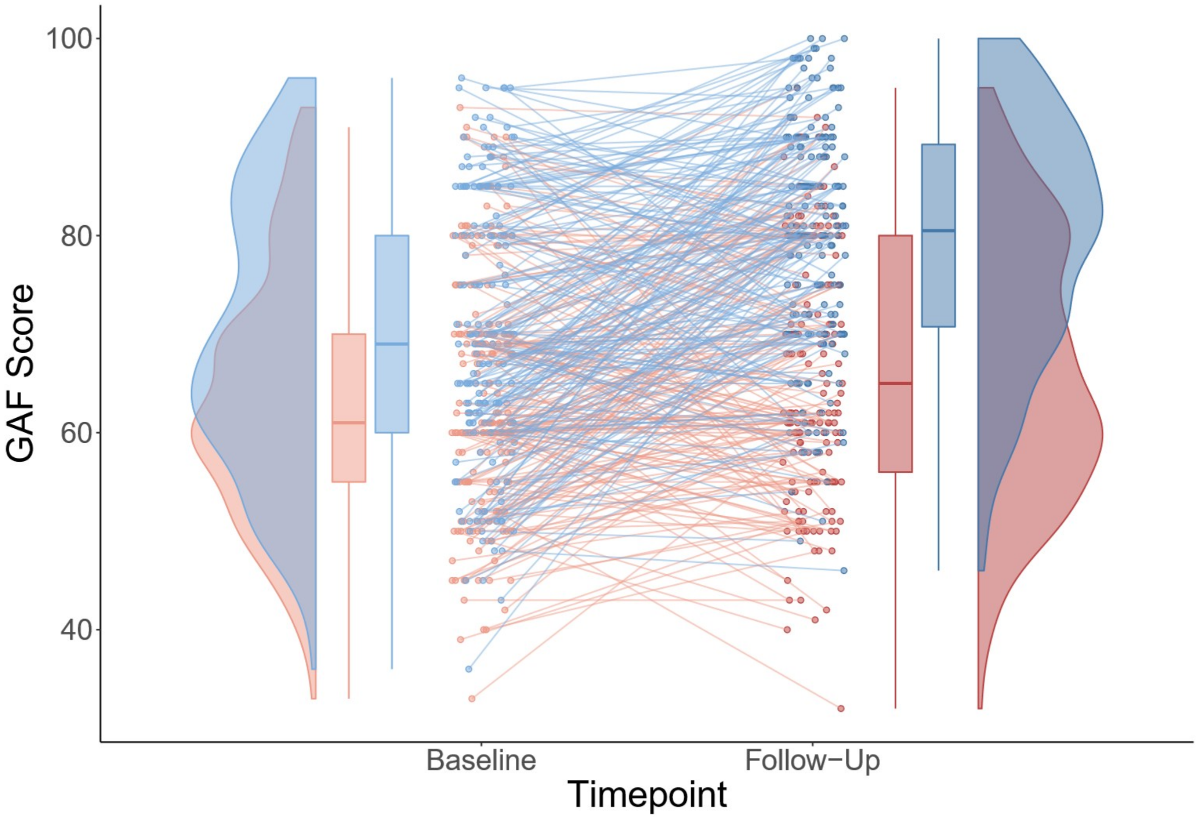


*Note.* Red = Relapse, Blue = No Relapse, points refer to individual scores, with lines connecting the two values of one person. For each timepoint, vertical boxplots and density distributions were added. Two participants from the Relapse group and three from the No Relapse group were excluded from this figure due to missing values in GAF score at baseline. The plot was created with the raincloudplot R package (van Langen, 2023).

#### Figure S4

*Permutation Distributions of the Relapse Models*

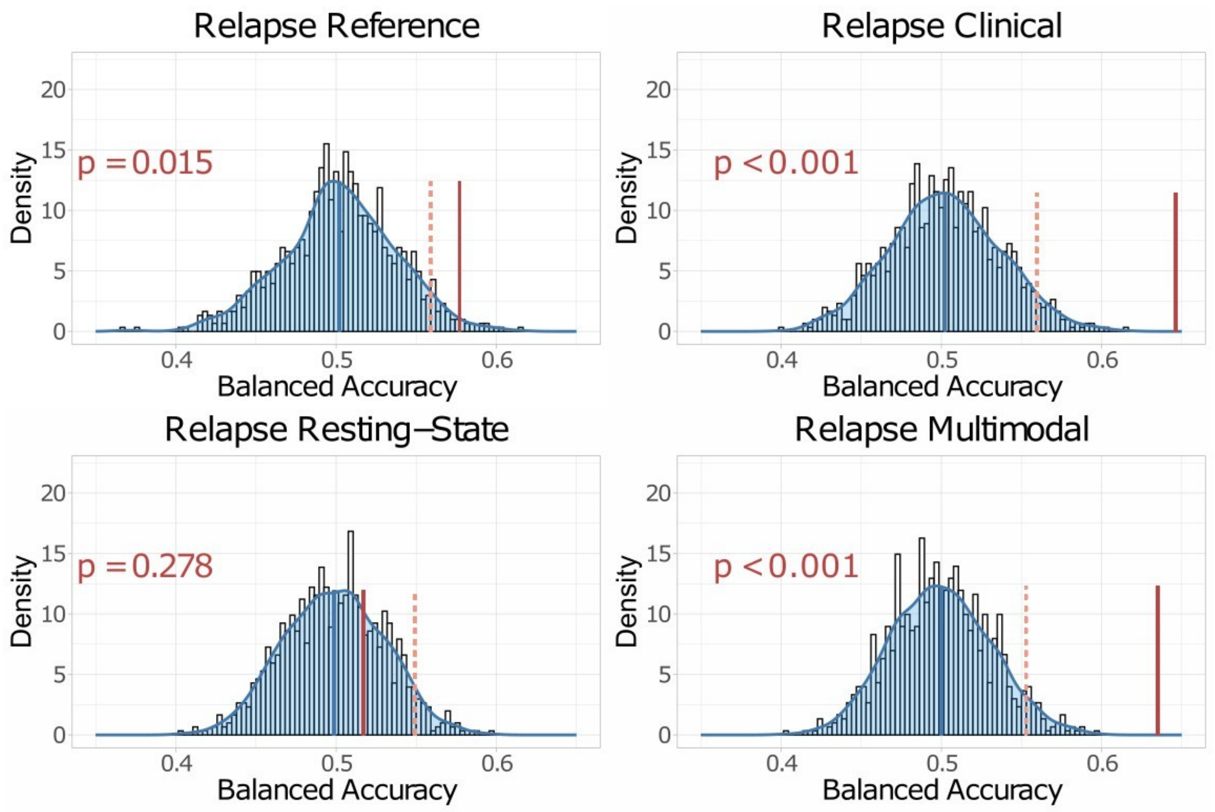


*Note.* The dashed red lines mark the 95% cutoffs of the permutation distributions, the solid red lines mark the observed balanced accuracies. Exact p-values are printed in red and indicate the portion of permutation results that are larger than the respective observed result.

### Figure S5

*Permutation Distributions of the GAF Models*

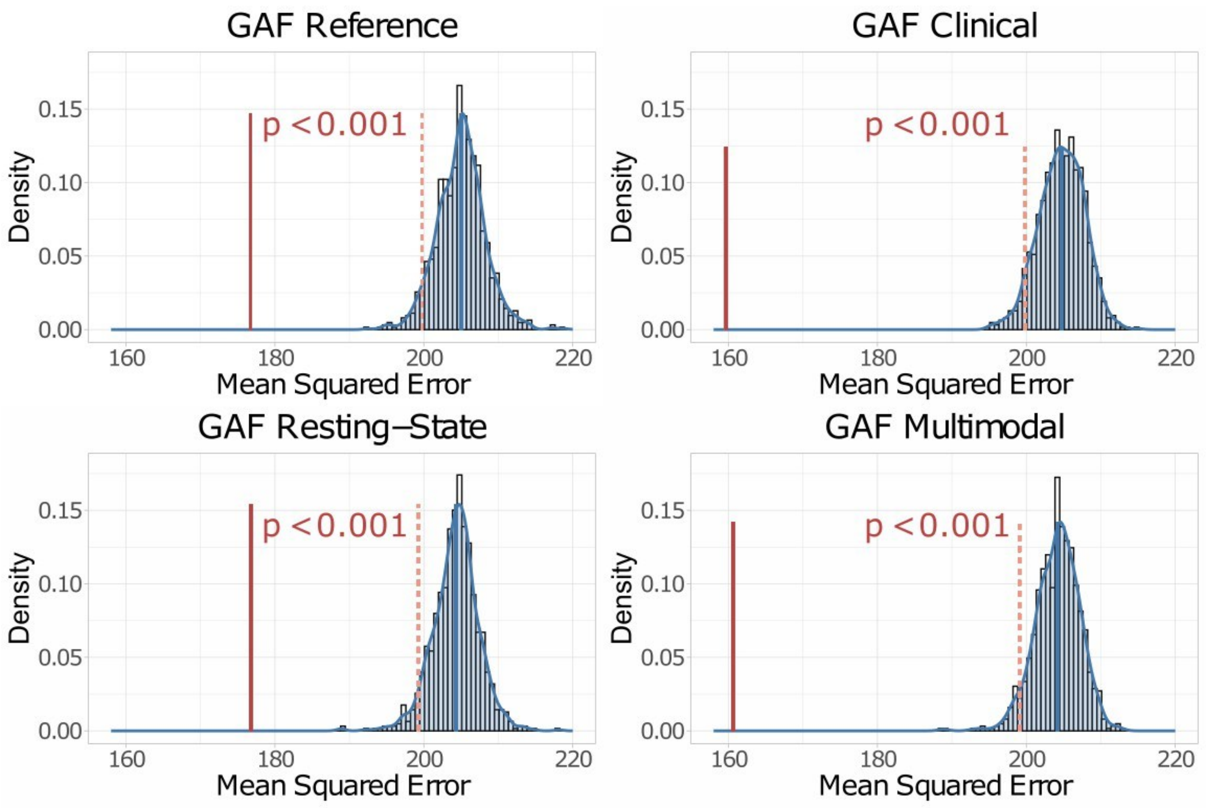


*Note.* The dashed red lines mark the 5% cutoffs of the permutation distributions, the solid red lines mark the observed balanced accuracies. Exact p-values are printed in red and indicate the portion of permutation results that are smaller than the respective observed result.

### Figure S6

*Comparisons of Classification Models for the Prediction of Relapse*

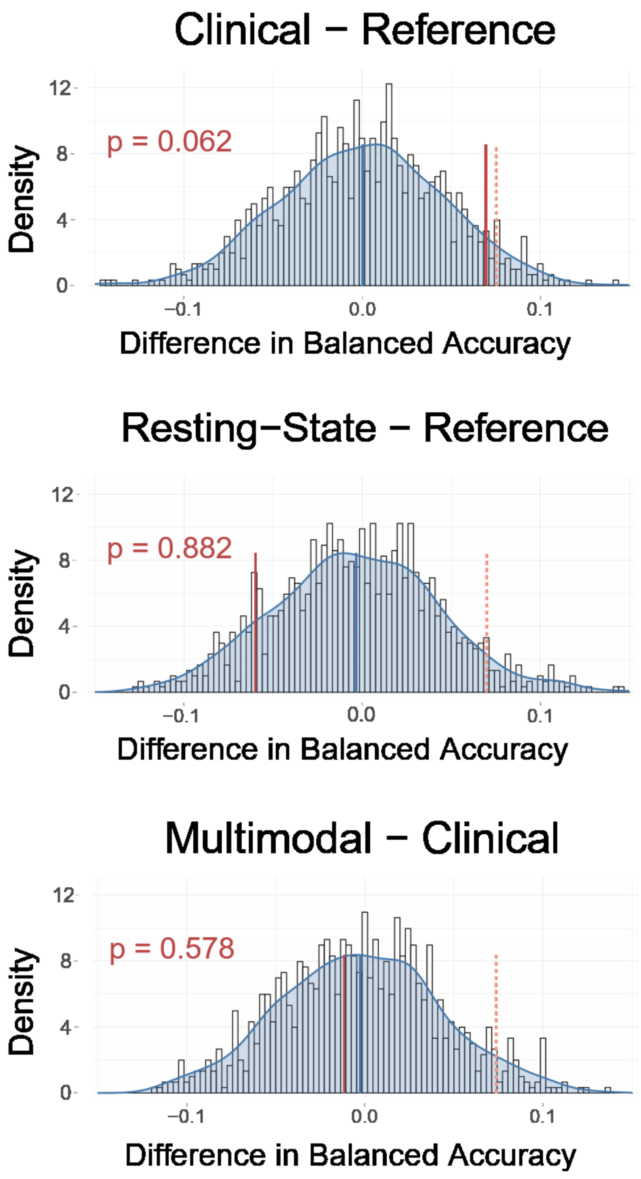


*Note.* The figure shows the distributions of the subtracted permutation performances from the classification models. The upper panel shows the test of the Clinical versus the Reference model, the mid panel of the Resting-State versus the Reference model, and the bottom panel of the Multimodal versus the Clinical model. Blue vertical lines are the means and red lines are the 95% cutoffs of the distributions, solid red lines are the observed differences. Exact p-values are annotated in red and indicate the portion of permutation differences that are larger than the respective observed result.

### Figure S7

*Comparisons of Regression Models for the Prediction of GAF*

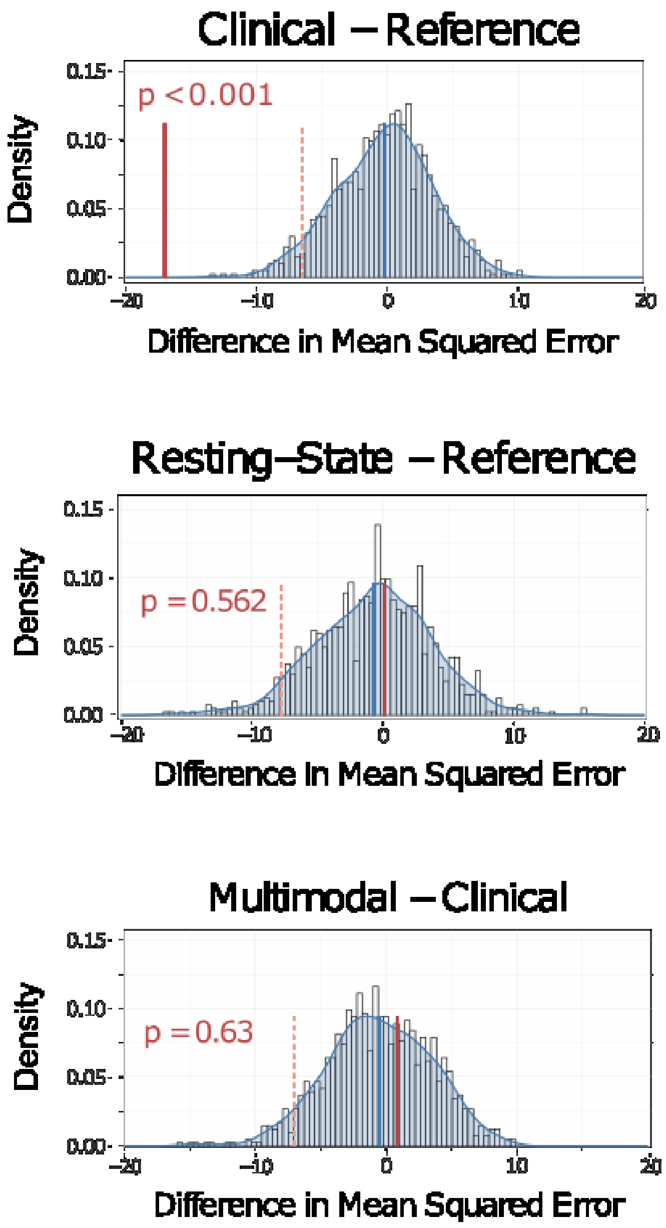


*Note.* The figure shows the distributions of the subtracted permutation performances from the regression models. The upper panel shows the test of the Clinical versus the Reference model, the mid panel of the Resting-State versus the Reference model, and the bottom panel of the Multimodal versus the Clinical model. Blue vertical lines are the means and red lines are the 95% cutoffs of the distributions, solid red lines are the observed differences. Exact p-values are annotated in red and indicate the portion of permutation differences that are larger than the respective observed result.

### Figure S8

*Mean Performance in the Training and Test Sets of the Relapse Models*

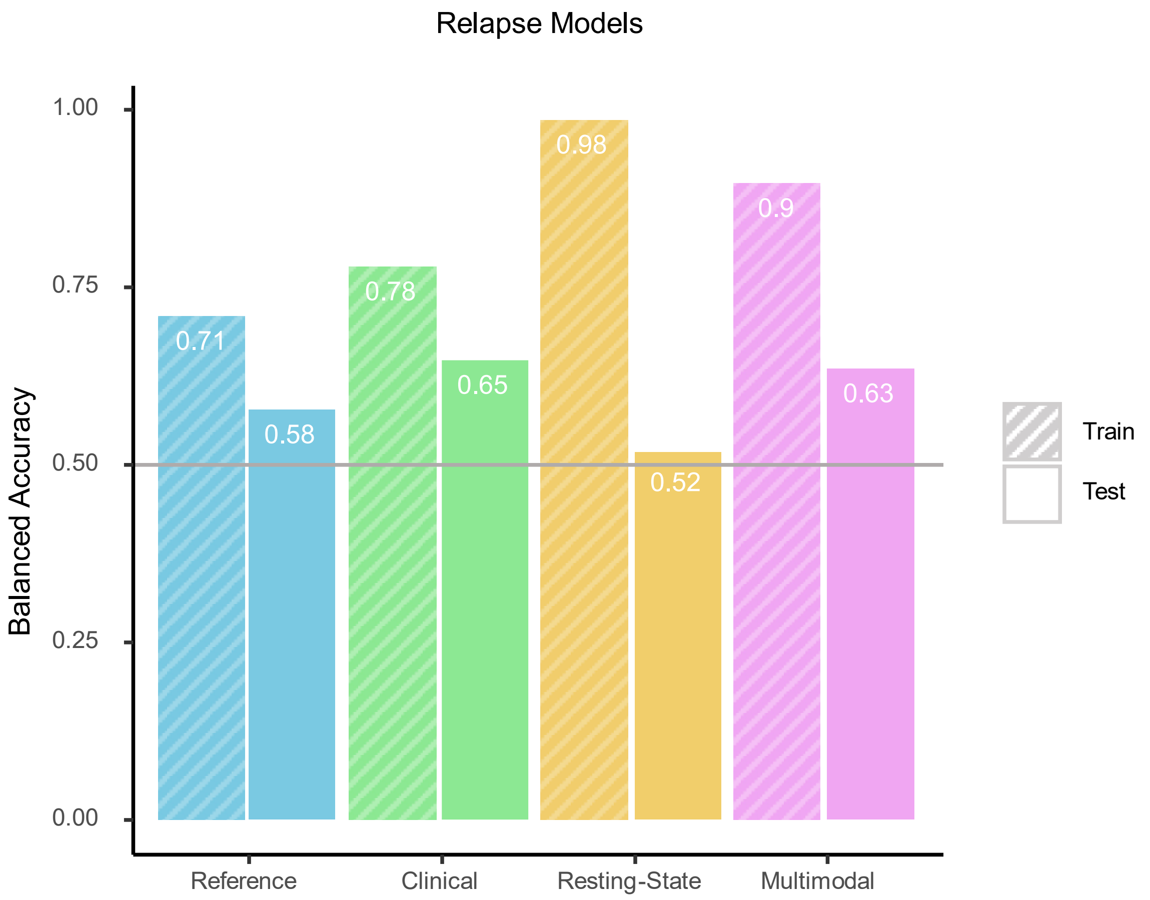


*Note.* The horizontal gray line marks the dummy performance which is chance level at 50% balanced accuracy. Larger balanced accuracy means better performance. Better performance in the training than the test sets implies overfitting of the model.

### Figure S9

*Mean Performance in the Training and Test Sets of the GAF Models*

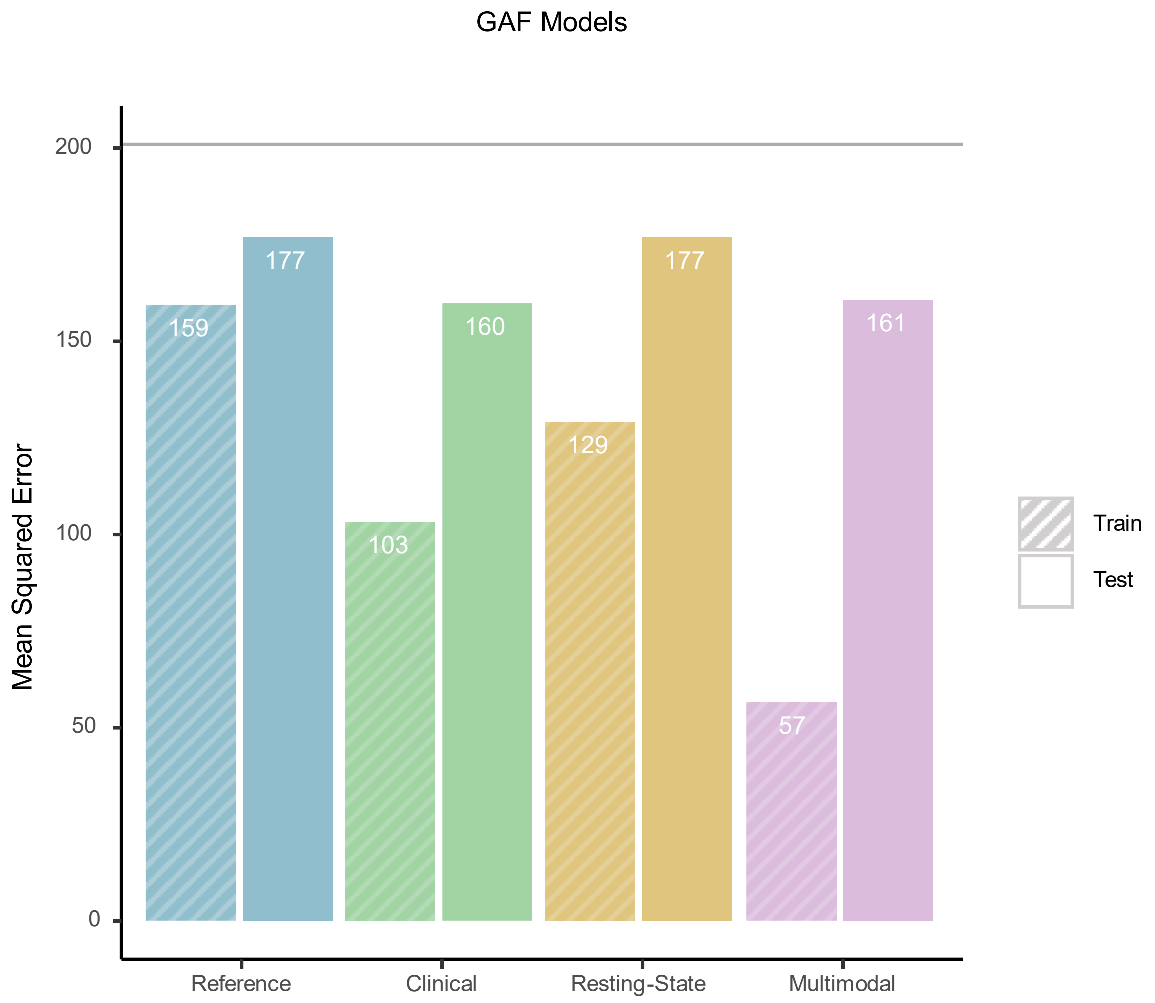


*Note.* The horizontal gray line marks the dummy performance which is chance level at 200.87 mean squared error. Smaller mean squared error means better performance. Better performance in the training than the test sets implies overfitting of the model.
